# Differences in Family Dementia Caregiver Needs and Preferences Across the Lifespan

**DOI:** 10.64898/2026.05.15.26353316

**Authors:** Virginia Gallagher, Christina Sheehan, Carol Manning, Kelly Shaffer

## Abstract

**Background:** The majority of family dementia caregivers in the United States (U.S.) are now young and middle-aged adults. However, little research has been conducted to understand how caregiver needs and preferences for support differ depending on their phase of adulthood. This study evaluated differences in mental health, caregiving readiness, desired supports, and intervention preferences among early (<46 years), middle (46–60 years), and late (>60 years) adulthood dementia caregivers.

**Methods:** A cross-sectional survey was conducted with 202 family dementia caregivers aged 22 to 88. Caregivers completed validated measures of burden, anxiety, depression, well-being, time pressure, dementia knowledge, caregiving preparedness, and positive aspects of caregiving. Desired supports and preferences for intervention format, program type, and frequency were assessed. Analyses examined both categorical adulthood phase and continuous age associations with caregiver outcomes, with alpha thresholds of p<.05.

**Results:** Early adulthood caregivers self-reported higher anxiety symptoms (relative to late adulthood caregivers) and perceived time pressure (relative to middle and late adulthood caregivers). Relative to late adulthood caregivers only, early adulthood caregivers more frequently endorsed desired support for supplemental care and safety tools for the person with dementia, as well as willingness to engage in individual counseling and automated, digital supports. Relative to both middle adulthood and late adulthood caregivers, they also more frequently expressed desired support for their own mental health.

**Conclusions:** Dementia caregiving in early adulthood is associated with distinct psychological and practical support needs, suggesting life course–informed interventions may enhance relevance and engagement.

**Key Points:**

- Interventions to support dementia caregiver mental health and burden have historically overlooked the quickly growing contingent of early to middle adulthood, non-spousal caregivers. This paper addresses a critical gap in our understanding of mental health, caregiving readiness, desired supports, and intervention preferences among these cohorts.
- Early adulthood dementia caregivers report greater anxiety, perceived time pressure, desire for mental health and supplemental care support, and willingness to engage in digital support interventions.
- Effective interventions for mental health and caregiving readiness among dementia caregivers should address distinct practical and psychological support needs and preferences across different phases of adulthood.

## Background and Objectives

There are nearly 12 million adults in the U.S. who provide unpaid care to a family member or friend with dementia due to Alzheimer’s and other neurodegenerative diseases(Alzheimer’s Association 2025). Approximately 70% of these dementia caregivers are young and middle-aged adults(Alzheimer’s Association 2025). Dementia caregivers are, on average, younger now than in previous decades due to sociodemographic trends such as older childbearing age (rendering adult children younger at time of parental ageing), smaller family sizes (fewer siblings to distribute care responsibilities), and more older adults ageing without spousal support (due to separation) (Freedman *et al*. 2023; Wolff *et al*. 2025). Despite this trend, the field has paid limited attention to the distinct needs and circumstances of younger and middle-aged dementia caregivers.

Regardless of age, dementia caregivers report elevated mental health distress relative to non-caregiving peers (Ma *et al*. 2018; Pinquart and Sörensen 2003). Hundreds of non-pharmacological interventions have been developed to mitigate this stress (Walter and Pinquart 2020); however, young and middle-aged caregivers have often been overlooked in these interventions. There is substantial overrepresentation of older, spousal caregivers in relevant research samples: although only about 10% of dementia caregivers are spousal caregivers, an average of 53% of dementia caregiver intervention trial participants are spousal, with a mean age of 62 (Alzheimer’s Association 2025; Walter and Pinquart 2020). This mismatch suggests that the needs, constraints, and preferences of young and middle-aged dementia caregivers remain insufficiently understood or addressed.

Age-related contextual factors, such as employment demands, parenting responsibilities, and digital technology proficiency, likely influence preferences for format, frequency, needed content, and ultimately intervention efficacy (Walter and Pinquart 2020). Understanding how caregiver needs differ by age or life phase requires a theoretical framework that captures common themes and role-conflicts that occur during different phases of adulthood.

The life course perspective offers a useful lens for examining how caregiver needs and intervention preferences vary by adulthood phase (Elder and George 2016). The framework describes the roles and responsibilities characteristic of different phases of adulthood and emphasizes how historical context shapes attitudes and behaviors. Although age cutoffs vary across studies, adulthood phases are often approximated as early adulthood representing ages ∼18–40, middle adulthood ages ∼41–59, and late adulthood ages 60+ (Hutteman *et al*. 2014). Individuals born in the same period often share similar life-cycle stages (e.g., midlife parent vs. retiree) as well as common attitudes, behaviors, and preferences influenced by major national events, economic recessions, and technological change (Geiger 2015). For example, caregivers who were aged 45 or younger when smartphones became ubiquitous in the U.S. in the 2010s are more likely to report using health-related smartphone apps and social media for health-related purposes compared to caregivers who were aged 45 or older during that period (Gallagher *et al*. 2024). Little research has directly compared dementia caregivers across adulthood phases, a problematic gap given that the limited available studies suggest potential group-based, non-linear associations between age and caregiver outcomes (Borsje *et al*. 2016).

Emerging evidence suggests that early and middle adulthood caregivers experience a distinct caregiving context relative to older caregivers. While caregiving demands are similar across caregiver adulthood phases, early and middle adulthood caregivers often have additional life-stage stressors such as childrearing, work demands, commuting, and disrupted education/career trajectories superimposed on caregiving stressors (Hou *et al*. 2016; Tong *et al*. 2026). In particular, compared to older adult family caregivers, younger caregivers more often report financial strain, likely related to ongoing income dependence and fewer accumulated resources (Koumoutzis *et al*. 2021; Wang *et al*. 2011). Many early and middle adulthood caregivers are “sandwiched caregivers,” in which they are simultaneously caring for an ageing parent with dementia while raising their own children (Lei *et al*. 2023). This dual role is associated with greater emotional caregiving burden, lower well-being due to increased work-family conflict, and cumulative stress exposure (Nemmers *et al*. 2025; Ory *et al*. 1999). Psychosocially, sandwich caregiving can produce emotional spillover across domains, where stress related to managing behavioral symptoms of dementia affects parenting quality and family functioning, while competing caregiving demands contribute to guilt, role overload, and reduced well-being (Lei *et al*. 2023; Nemmers *et al*. 2025). In contrast, older caregivers are more likely to experience burden associated with physical caregiving demands and age-related health limitations rather than competing role obligations (Schulz and Sherwood 2008). Despite advances in understanding of caregiver contexts across the lifespan, little work has been done to rigorously explore differences in caregiver mental health, readiness, and intervention preferences cross-sectionally across dementia caregiver age-groups.

The present study evaluates the hypothesis that mental health symptoms, caregiving readiness, caregiver support needs, and caregiver intervention preferences differ across adulthood phases among dementia caregivers. By identifying potential life stage–related patterns, this work aims to inform the refinement of existing interventions and the design of future tailored interventions to best meet the needs of dementia caregivers across each of the adulthood phases.

## Research Design and Methods

### Study Design and Participants

This study employed cross-sectional surveys to evaluate patterns of caregiving mental health symptoms, support needs, and intervention preferences among adults at different phases of adulthood. The analytic sample comprises 202 adults who provide unpaid care to an older adult with cognitive changes impacting independence in activities of daily living (henceforth, “caregivers”). The analytic sample included n=87 caregivers from a study that recruited caregivers in Virginia only (Study A) and n=115 caregivers from an online study that utilized national recruitment methods (Study B). Both studies were approved by the site’s Institutional Review Board. In both studies, eligible individuals were aged 18 or older, able to read and write in English, and actively assisting an individual aged 55 or older in need of support due to cognitive changes that impact their day-to-day activity. Individuals were required to provide caregiving support for at least two activities of daily living over the past three months without receiving payment (with the exception of Medicaid family caregiver payment). Individuals were ineligible to participate if they themselves had a diagnosis of cognitive impairment, or if their self-rated Eight-item Informant Interview to Differentiate Aging and Dementia (AD8) score was less than two, suggesting a possible absence of cognitive impairment in the care-recipient. The only difference in inclusion/exclusion criteria between the two studies was that Study A required the care-recipient of the caregiver to live in the community (i.e., not in a continuing care facility), whereas in Study B, there were no restrictions on care-recipient living environment. To support the examination of adulthood phase and its impacts on caregiver outcomes, participants younger than 60 were oversampled. Once the enrollment goal (n=85) was met for participants older than 60, enrollment for that cohort ceased, while enrollment of younger participants continued. This process allowed for greater representation in group analyses (116 early and middle adulthood caregivers (n=59 and n=57, respectively) versus 86 late adulthood caregivers).

## Procedures

### Recruitment and Screening

Caregivers were recruited from a Memory Disorders Clinic at an academic medical center (n=81, 40.1%), online advertising via social media, websites, and email list-serves (n=59, 29.2%), by word-of-mouth (n=17, 8.4%), community events (n=11, 5.5%), physical flyers posted in Virginia (n=11, 5.5%), or “other” (n=17, 8.4%). Recruitment method was self-reported by participants and cross-referenced with study recruitment logs; recruitment method was missing for n=6 participants (3%). Interested caregivers were invited to complete an online screening form in REDCap or by phone. Eligible caregivers were screened for fraud in a multi-step process guided by previously published literature (Davies *et al*. 2024; Glazer *et al*. 2021); see Supplementary Material for more details. Eligible, validated individuals either completed an informed consent process by phone with a study staff member and signed a digital consent form (Study A) or reviewed an information sheet about the study outlining the activities, risks, and benefits with consent implied by returning surveys (Study B).

### Data Collection and Payment

Individuals who proceeded to study activities completed a series of questionnaires online that required an estimated 45 minutes to complete. They were invited to complete them at their own pace but within 30 days of questionnaire receipt. Participants who expressed difficulty with digital tools could complete the questionnaires by phone or by paper. Participants who completed surveys in their entirety for Study A were sent an electronic $35 gift card and an additional $50 payment for an elective blood draw procedure. Participants who completed surveys in their entirety for Study B were sent an electronic $85 gift card.

## Measures

### Demographic and Caregiving Characteristics

Demographic and caregiving characteristic variables originated from caregivers’ responses to a novel Demographics and Caregiving Form, which was composed from items from the Uniform Data Set – version 4.0 Informant Questionnaire, the BRFSS Caregiving Module 2019, and study-team generated items that were iterated via four group discussions and revisions (blank form provided in Supplementary Materials).

Self-reported age on the Demographics and Caregiving Form was used to divide caregivers into early (age < 46 years), middle (age 46-60 years), and late (age > 60 years) adulthood phases for analysis. Age cut-offs for adulthood phase are variable in the U.S. public health literature, with cut-off ages for middle adulthood ranging from 40 to 45 (on the early end) and 60 to 65 (on the late end) (Halloran 2024; Kalmijn *et al*. 2004; Pope *et al*. 2022; Zahodne *et al*. 2017), with groupings a best attempt to reflect common conflicts, demands, and goals that often happen during specific age bands (Halloran 2024).

### Caregiver Mental Health and Caregiving Readiness

Caregiving emotional burden was assessed with the 12-item Zarit Burden Interview – Short Form (ZBI-SF), with higher sum scores reflecting higher burden (sample Chronbach’s α = .913). Anxiety symptoms were measured using the 7-item Anxiety Subscale of the Hospital Anxiety and Depression Scale (HADS-Anxiety), with higher sum scores reflecting higher anxiety (α = .836). Depressive symptoms were measured using the 7-item Depression Subscale of the Hospital Anxiety and Depression Scale (HADS-Depression), with higher scores representing higher depression (α = .823). Well-being was assessed with the 5-item World Health Organization-Five Well-Being Index (WHO-5), with higher scores indicating greater well-being (α = .888). Time pressure was measured using Garhammer’s 7-item Index of Time Pressure, with higher scores reflecting greater time pressure (α = .910). Dementia knowledge was assessed with the 21-item Dementia Knowledge Assessment Tool Version Two (DKAT-2), with higher scores reflecting greater percentage of correctly answered items (α = .791). Caregiving preparedness was measured using the 8-item Preparedness for Caregiving Scale (PCS), with higher scores reflecting greater perceived preparedness (α = .894). Caregiving positivity was assessed with the 6-item Positive Aspects of Caregiving Scale (PACS), with higher scores indicating higher positivity about caregiving (α = .891).

### Caregiver Desired Supports and Preferences for Intervention

Caregivers’ areas of desired support were assessed by answers to the following question: “If we could wave a magic wand and provide support for you in your role as a caregiver for the person with cognitive decline, what would you want help with?” Caregivers could choose any number of desired supports from eight categories (caregiver mental health, caregiving skills, caregiver wellness, medical advice related to the person with dementia [PWD], advanced care planning, supplemental PWD care, PWD safety, and PWD wellness) with two to five support options within each category.

Caregivers’ intervention preferences were assessed by four questions about preferred support setting (choose one: in-person, online, hybrid, other), program type (select all that apply: individual counseling, support group, podcasts/videos/self-guided mobile app, other), and support frequency (choose one: weekly, monthly, other).

## Data Analysis

Information regarding data cleaning is in the Supplemental Material. Demographic and caregiving characteristics were summarized descriptively. The primary hypothesis was tested with group comparisons to identify potential associations between adulthood phase and a) caregiver mental health and readiness measures via analyses of variance and b) caregiver needs and preferences for intervention via chi-square analyses. Post-hoc pairwise group comparisons were conducted when main effect tests were significant. Follow-up analyses were conducted using age as a continuous variable and the same dependent variables to evaluate the extent to which the relationship between age and variables of interest were linear in nature versus categorical. These analyses involved identifying associations between age and a) caregiver mental health and readiness measures via Pearson’s correlations and b) caregiver needs and preferences for intervention via t-tests comparing mean age of those who endorsed versus did not endorse identified needs and preferences. For continuous outcome variables, when assumptions of normality (Shapiro-Wilk tests) or homogeneity of variance (Levene’s test) were not met, Mann-Whitney U or Kruskal-Wallis tests were used for two and three group comparisons, respectively. Analyses were conducted with IBM SPSS Statistics 28. All tests were two-sided, with *p*-values < .05 interpreted.

## Results

The sample included 202 dementia caregivers across early adulthood (n=59), middle adulthood (n=57), and late adulthood (n=86). The majority of caregivers were women (n=163, 80.7%), White (n=177, 87.6%), at least college educated (n=159, 79.1%), married/partnered (n=157, 77.7%), and working for pay (n=33, 16.3% working 1-20 hours per week; n=92, 45.5% working 21 hours or more per week) (see Table 1). Regarding caregiving characteristics, 47 (23.3%) caregivers also reported having another care-recipient with a chronic condition. Caregivers were most often the spouse (n=89, 44.1%) or the child of the care-recipient (n=78, 38.6%) with cognitive impairment, and over two-thirds of caregivers (n=137, 67.8%) lived with the care-recipient. The vast majority (n=179, 88.6%) had provided care to the person with cognitive decline for at least one year. Care intensity (hours per week) varied across the sample (see Table 2). While not evaluated statistically, demographic and caregiving trends within caregiver adulthood phases mirrored what has been previously demonstrated in national caregiving samples (Gallagher *et al*. 2025).

**Table 1.**
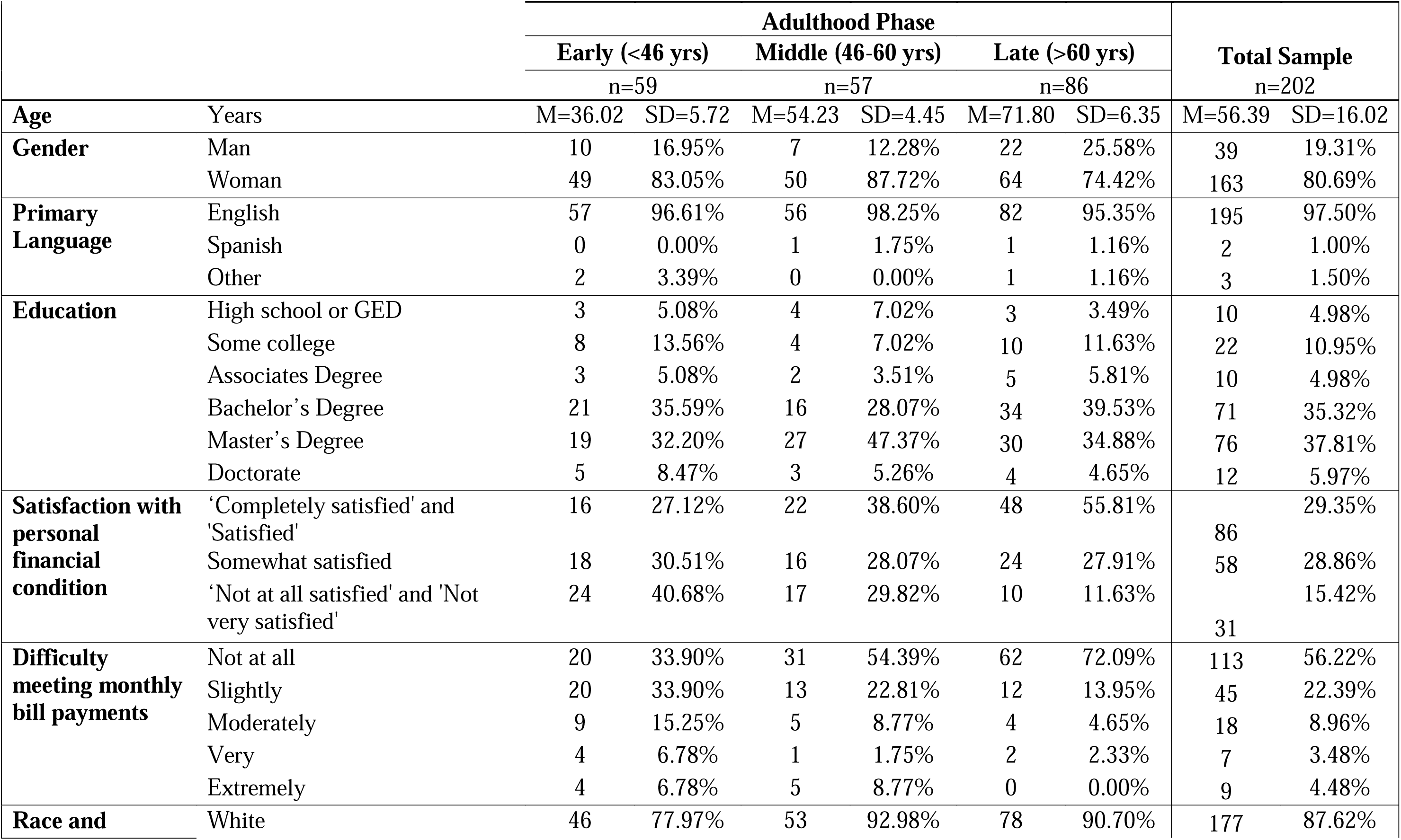
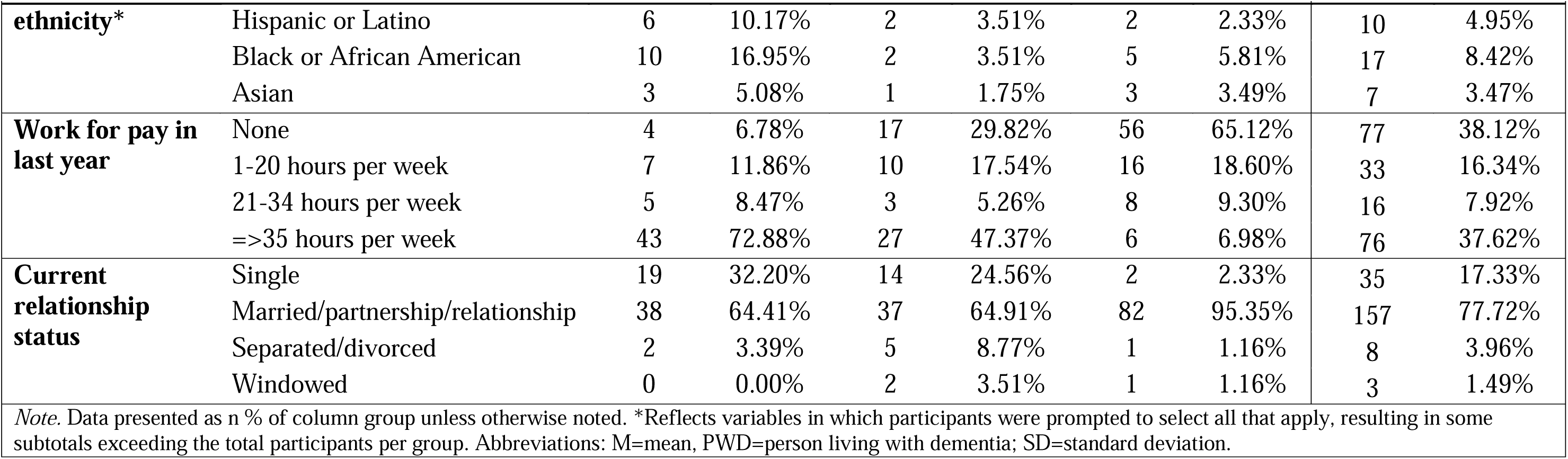
Demographic Characteristics.

**Table 2.**
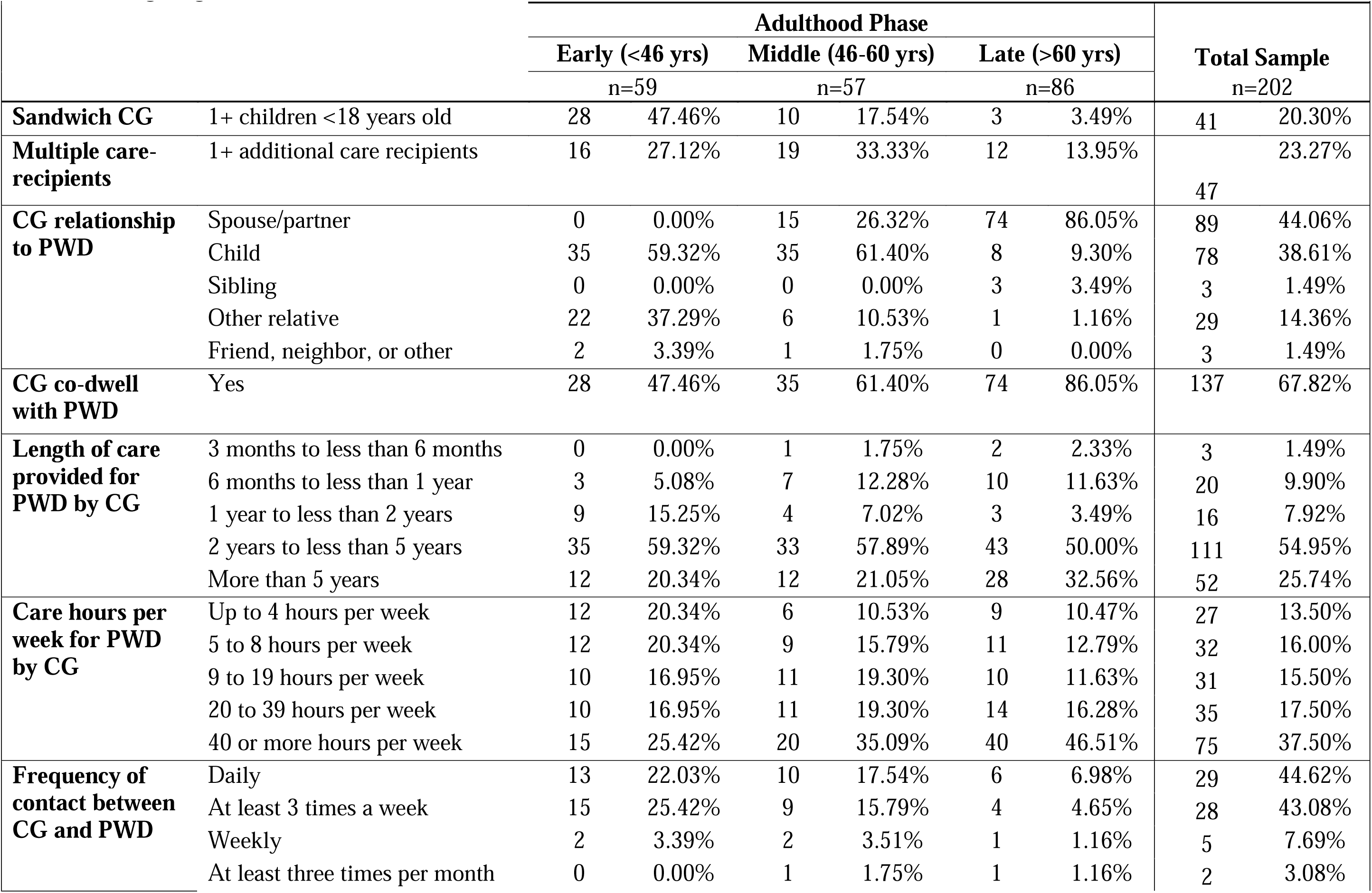
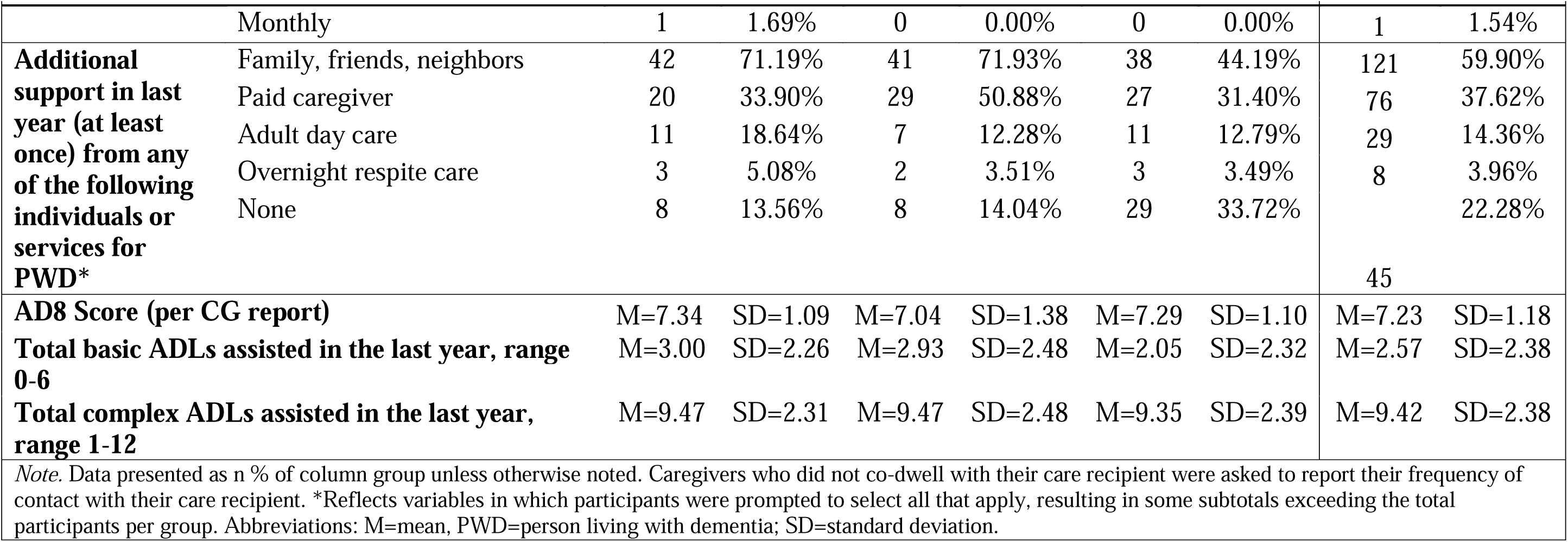
Caregiving Characteristics.

### Mental Health and Caregiving Readiness

Across the entire sample, mean emotional caregiving burden fell in the high burden range (>20) (Higginson *et al*. 2010), mean depression symptoms within the normal range (≤ 7) (Wu *et al*. 2021), mean anxiety symptoms in the significant range (≥8) (Olssøn *et al*. 2005; Puhan *et al*. 2008), and mean well-being in the poor range (<13) (Topp *et al*. 2015). Additionally, sample mean dementia knowledge scores indicated an average of 66.7% of items were answered correctly, and mean preparedness for caregiving scale indicated that on average, caregivers felt “somewhat well prepared” for various aspects of caregiving. There are no interpretation guidelines for the preparedness measure.

### Caregiver Adulthood Phase Analyses

#### Categorical Adulthood Phase Analyses

There was a main effect of adulthood phase on anxiety symptoms [F (2,199) = 4.41, p = .013] and time pressure [F (2, 193) = 9.95, p < .001] but not on caregiver burden, depression symptoms, well-being, dementia knowledge, positive aspects of caregiving, or preparedness for caregiving. Post-hoc testing revealed that early adulthood caregivers had significantly higher self-reported symptoms of anxiety relative to middle and late adulthood caregivers, and middle adulthood caregivers significantly higher than late adulthood caregivers (see Table 3). Further, time pressure was significantly higher among early adulthood caregivers relative to older adulthood caregivers.

**Table 3.** Caregiver Mental Health and Readiness by Adulthood Phase and Age.

|  |  |  |  |  |  |  |  |  |  |
| --- | --- | --- | --- | --- | --- | --- | --- | --- | --- |
| *Caregiver construct* | **Adulthood Phase Analyses** |  |  |  |  |  |  |  | **Continuous Age Analyses** |
|  | **Early (<46 yrs)** |  | **Middle (46-60 yrs)** |  | **Late (>60 yrs)** |  | **Total Sample** |  | **Age** |
|  | *Mean* | *SD* | *Mean* | *SD* | *Mean* | *SD* | *Mean* | *SD* | *Pearson's r* |
| **Caregiver Emotional Burden** | 24.75 | 8.98 | 23.37 | 9.43 | 21.79 | 9.81 | 23.10 | 9.50 | -.174* |
| **Depression Symptoms<sup>^</sup>** | 6.69 | 3.32 | 6.23 | 3.94 | 5.80 | 3.71 | 6.18 | 3.67 | -.157* |
| **Anxiety Symptoms** | 9.69<sub>a</sub> | 4.79 | 8.42<sub>a,b</sub> | 3.95 | 7.55<sub>b</sub> | 4.11 | 8.421 | 4.35 | -.239** |
| **Quality of Life** | 12.34 | 5.21 | 12.21 | 5.41 | 13.09 | 5.72 | 12.63 | 5.48 | .109 |
| **Time Pressure** | 23.73<sub>a</sub> | 6.43 | 20.95<sub>b</sub> | 6.12 | 18.95<sub>b</sub> | 6.23 | 20.96 | 6.54 | -.347*** |
| **Dementia Knowledge<sup>^</sup>** | 14.56 | 3.77 | 14.75 | 3.63 | 14.03 | 3.46 | 14.39 | 3.60 | -.085 |
| **Positive Aspects of Caregiving<sup>^</sup>** | 19.78 | 5.94 | 19.46 | 5.85 | 19.95 | 5.82 | 19.76 | 5.83 | .016 |
| **Preparedness for Caregiving** | 18.52 | 5.25 | 18.41 | 6.40 | 17.91 | 6.70 | 18.23 | 6.20 | .002 |
| *Note.* <u>Categorical adulthood phase analyses:</u> Constructs that are shaded indicate significant main effects of adulthood phase on the relevant measure. <sup>^</sup> indicates that Kruskal-Wallis test was used due to assumption violation; otherwise, ANOVA was used. Results of post-hoc analyses are represented by subscript letters; values in the same row not sharing the same subscript are significantly different at $p < .05$ in the two-sided test of equality for column means. <u>Continuous age analyses:</u> significant correlations represented by *$p < .05$, **$p < .01$, ***$p < .001$. Highlighted cells indicate significant differences. |  |  |  |  |  |  |  |  |  |

#### Continuous Age Analyses

When age was examined as a continuous variable in this context, there were significant correlations such that younger age was associated with higher anxiety and time-pressure, and to a lesser degree, caregiver emotional burden and depression (see Table 3). There were no significant correlations between age and quality of life, dementia knowledge, positive aspects of caregiving, nor preparedness for caregiving

## Desired Supports

### Overall Sample

At least 70% of caregivers endorsed wanting support in every category assessed (see Table 4). The most frequently endorsed categories of desired support across the entire sample were caregiver mental health support (n=179, 88.6%), PWD wellness (n=172, 85.2%), caregiver skills training (n=166, 82.2%), and advanced care planning (n=163, 80.7%).

**Table 4.**
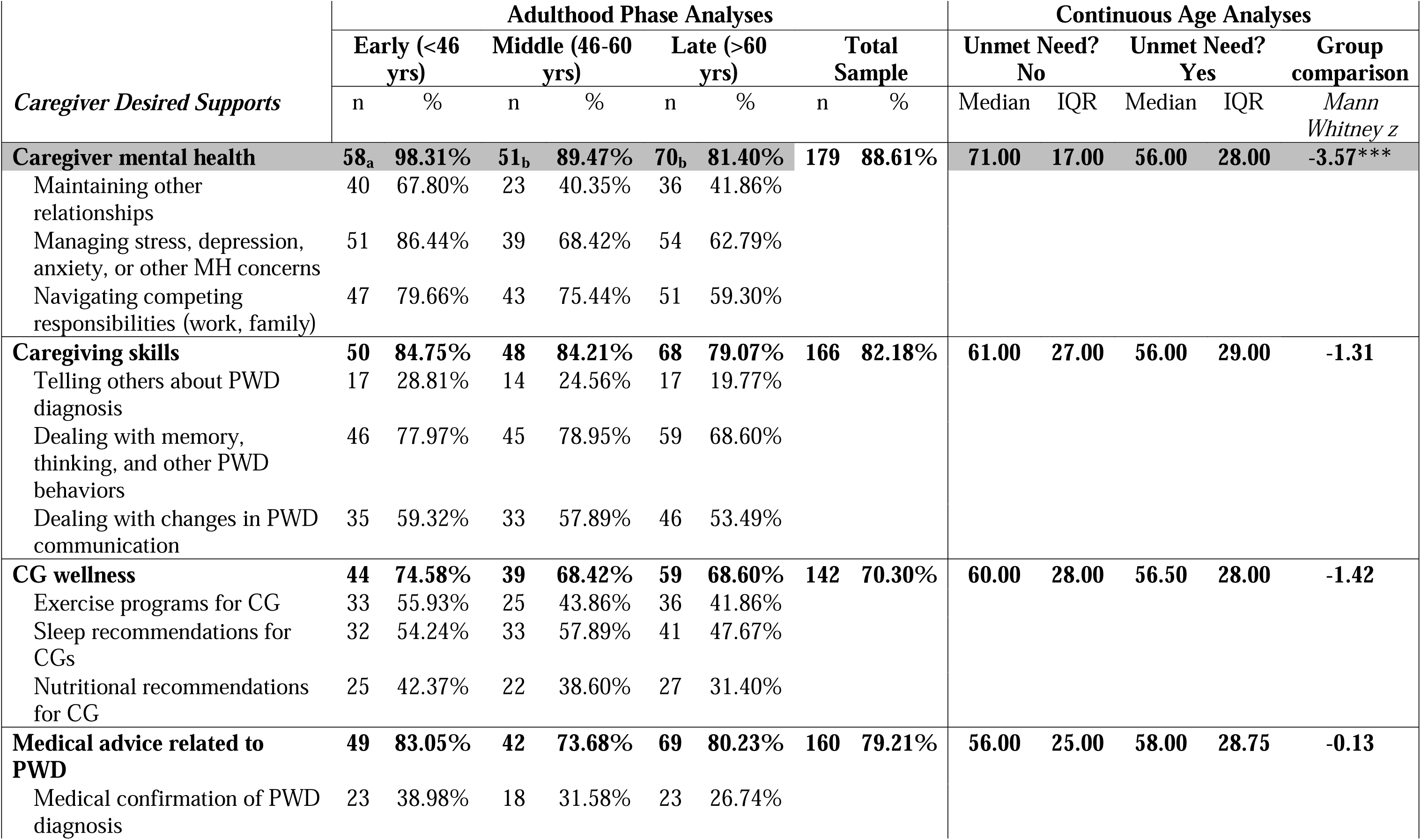
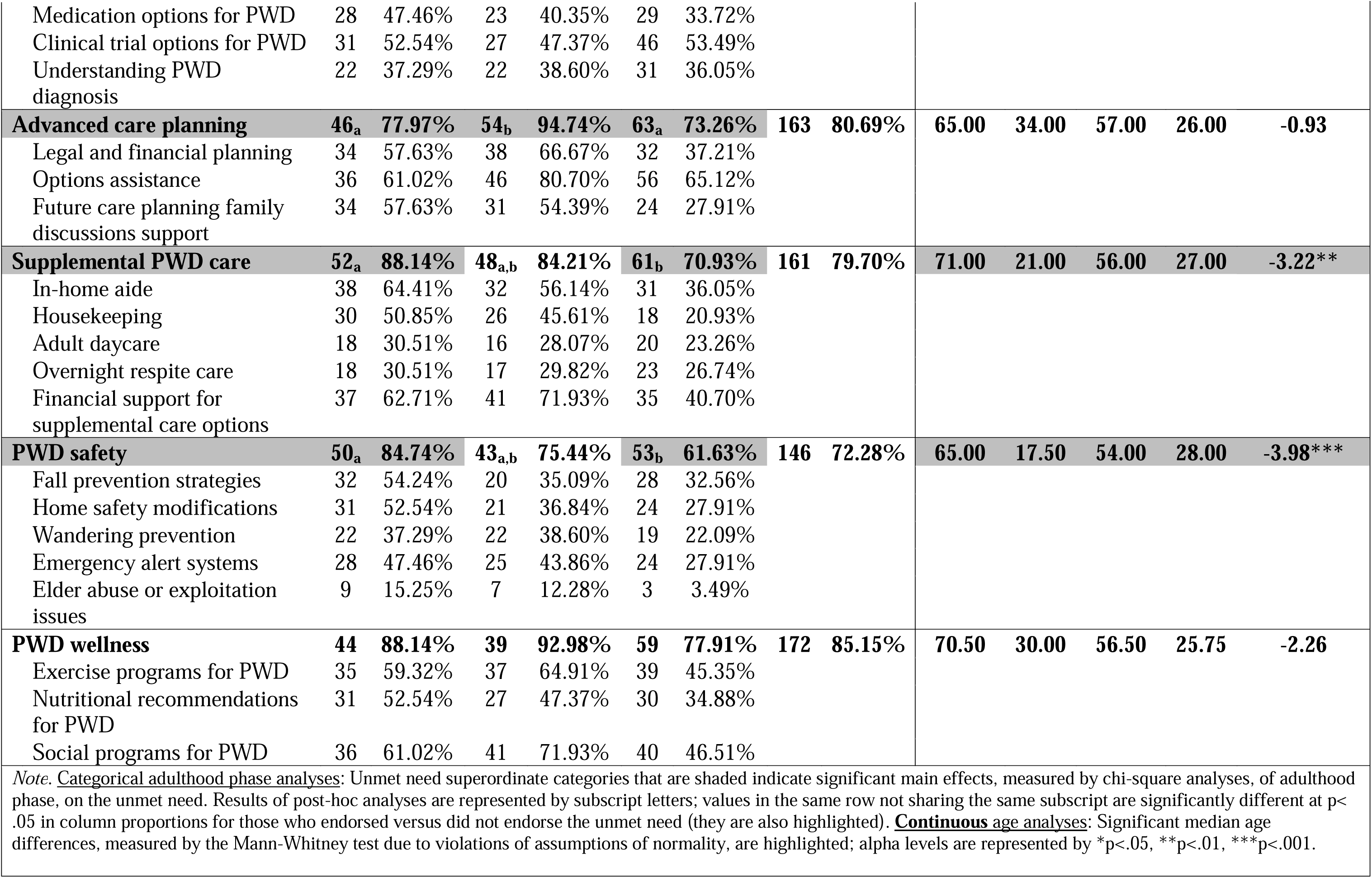
Caregiver Desired Supports by Adulthood Phase and Age.

| *Caregiver Desired Supports* | Adulthood Phase Analyses |  |  |  |  |  |  |  | Continuous Age Analyses |  |  |  |  |
| --- | --- | --- | --- | --- | --- | --- | --- | --- | --- | --- | --- | --- | --- |
|  | Early (<46 yrs) |  | Middle (46-60 yrs) |  | Late (>60 yrs) |  | Total Sample |  | Unmet Need? No |  | Unmet Need? Yes |  | Group comparison |
|  | n | % | n | % | n | % | n | % | Median | IQR | Median | IQR | *Mann Whitney z* |
| **Caregiver mental health** | **58<sub>a</sub>** | **98.31%** | **51<sub>b</sub>** | **89.47%** | **70<sub>b</sub>** | **81.40%** | **179** | **88.61%** | **71.00** | **17.00** | **56.00** | **28.00** | **-3.57\*\*\*** |
| Maintaining other relationships | 40 | 67.80% | 23 | 40.35% | 36 | 41.86% |  |  |  |  |  |  |  |
| Managing stress, depression, anxiety, or other MH concerns | 51 | 86.44% | 39 | 68.42% | 54 | 62.79% |  |  |  |  |  |  |  |
| Navigating competing responsibilities (work, family) | 47 | 79.66% | 43 | 75.44% | 51 | 59.30% |  |  |  |  |  |  |  |
| **Caregiving skills** | **50** | **84.75%** | **48** | **84.21%** | **68** | **79.07%** | **166** | **82.18%** | **61.00** | **27.00** | **56.00** | **29.00** | **-1.31** |
| Telling others about PWD diagnosis | 17 | 28.81% | 14 | 24.56% | 17 | 19.77% |  |  |  |  |  |  |  |
| Dealing with memory, thinking, and other PWD behaviors | 46 | 77.97% | 45 | 78.95% | 59 | 68.60% |  |  |  |  |  |  |  |
| Dealing with changes in PWD communication | 35 | 59.32% | 33 | 57.89% | 46 | 53.49% |  |  |  |  |  |  |  |
| **CG wellness** | **44** | **74.58%** | **39** | **68.42%** | **59** | **68.60%** | **142** | **70.30%** | **60.00** | **28.00** | **56.50** | **28.00** | **-1.42** |
| Exercise programs for CG | 33 | 55.93% | 25 | 43.86% | 36 | 41.86% |  |  |  |  |  |  |  |
| Sleep recommendations for CGs | 32 | 54.24% | 33 | 57.89% | 41 | 47.67% |  |  |  |  |  |  |  |
| Nutritional recommendations for CG | 25 | 42.37% | 22 | 38.60% | 27 | 31.40% |  |  |  |  |  |  |  |
| **Medical advice related to PWD** | **49** | **83.05%** | **42** | **73.68%** | **69** | **80.23%** | **160** | **79.21%** | **56.00** | **25.00** | **58.00** | **28.75** | **-0.13** |
| Medical confirmation of PWD diagnosis | 23 | 38.98% | 18 | 31.58% | 23 | 26.74% |  |  |  |  |  |  |  |

|  |  |  |  |  |  |  |  |  |  |  |  |  |  |  |
| --- | --- | --- | --- | --- | --- | --- | --- | --- | --- | --- | --- | --- | --- | --- |
| Medication options for PWD | 28 | 47.46% | 23 | 40.35% | 29 | 33.72% |  |  |  |  |  |  |  |  |
| Clinical trial options for PWD | 31 | 52.54% | 27 | 47.37% | 46 | 53.49% |  |  |  |  |  |  |  |  |
| Understanding PWD diagnosis | 22 | 37.29% | 22 | 38.60% | 31 | 36.05% |  |  |  |  |  |  |  |  |
| **Advanced care planning** | **46<sub>a</sub>** | **77.97%** | **54<sub>b</sub>** | **94.74%** | **63<sub>a</sub>** | **73.26%** | **163** | **80.69%** | **65.00** | **34.00** | **57.00** | **26.00** | **-0.93** |  |
| Legal and financial planning | 34 | 57.63% | 38 | 66.67% | 32 | 37.21% |  |  |  |  |  |  |  |  |
| Options assistance | 36 | 61.02% | 46 | 80.70% | 56 | 65.12% |  |  |  |  |  |  |  |  |
| Future care planning family discussions support | 34 | 57.63% | 31 | 54.39% | 24 | 27.91% |  |  |  |  |  |  |  |  |
| **Supplemental PWD care** | **52<sub>a</sub>** | **88.14%** | **48<sub>a,b</sub>** | **84.21%** | **61<sub>b</sub>** | **70.93%** | **161** | **79.70%** | **71.00** | **21.00** | **56.00** | **27.00** | **-3.22\*\*** |  |
| In-home aide | 38 | 64.41% | 32 | 56.14% | 31 | 36.05% |  |  |  |  |  |  |  |  |
| Housekeeping | 30 | 50.85% | 26 | 45.61% | 18 | 20.93% |  |  |  |  |  |  |  |  |
| Adult daycare | 18 | 30.51% | 16 | 28.07% | 20 | 23.26% |  |  |  |  |  |  |  |  |
| Overnight respite care | 18 | 30.51% | 17 | 29.82% | 23 | 26.74% |  |  |  |  |  |  |  |  |
| Financial support for supplemental care options | 37 | 62.71% | 41 | 71.93% | 35 | 40.70% |  |  |  |  |  |  |  |  |
| **PWD safety** | **50<sub>a</sub>** | **84.74%** | **43<sub>a,b</sub>** | **75.44%** | **53<sub>b</sub>** | **61.63%** | **146** | **72.28%** | **65.00** | **17.50** | **54.00** | **28.00** | **-3.98\*\*\*** |  |
| Fall prevention strategies | 32 | 54.24% | 20 | 35.09% | 28 | 32.56% |  |  |  |  |  |  |  |  |
| Home safety modifications | 31 | 52.54% | 21 | 36.84% | 24 | 27.91% |  |  |  |  |  |  |  |  |
| Wandering prevention | 22 | 37.29% | 22 | 38.60% | 19 | 22.09% |  |  |  |  |  |  |  |  |
| Emergency alert systems | 28 | 47.46% | 25 | 43.86% | 24 | 27.91% |  |  |  |  |  |  |  |  |
| Elder abuse or exploitation issues | 9 | 15.25% | 7 | 12.28% | 3 | 3.49% |  |  |  |  |  |  |  |  |
| **PWD wellness** | **44** | **88.14%** | **39** | **92.98%** | **59** | **77.91%** | **172** | **85.15%** | **70.50** | **30.00** | **56.50** | **25.75** | **-2.26** |  |
| Exercise programs for PWD | 35 | 59.32% | 37 | 64.91% | 39 | 45.35% |  |  |  |  |  |  |  |  |
| Nutritional recommendations for PWD | 31 | 52.54% | 27 | 47.37% | 30 | 34.88% |  |  |  |  |  |  |  |  |
| Social programs for PWD | 36 | 61.02% | 41 | 71.93% | 40 | 46.51% |  |  |  |  |  |  |  |  |
| *Note. Categorical adulthood phase analyses:* Unmet need superordinate categories that are shaded indicate significant main effects, measured by chi-square analyses, of adulthood phase, on the unmet need. Results of post-hoc analyses are represented by subscript letters; values in the same row not sharing the same subscript are significantly different at $p < .05$ in column proportions for those who endorsed versus did not endorse the unmet need (they are also highlighted). *Continuous age analyses:* Significant median age differences, measured by the Mann-Whitney test due to violations of assumptions of normality, are highlighted; alpha levels are represented by *$p < .05$, **$p < .01$, ***$p < .001$. | | | | | | | | | | | | | | |

### Caregiver Adulthood Phase Analyses

#### Categorical Adulthood Phase Analyses

The following desired support categories significantly differed by adulthood phase: caregiver mental health support [Χ^2^ (2, *N =* 202) = 9.975, p=.007], advanced care planning [Χ^2^ (2, *N =* 202) = 10.551, p=.005], supplemental PWD care [Χ^2^ (2, *N =* 202) = 7.401, p=.025], PWD safety support [Χ^2^ (2, *N =* 202) = 9.729, p=.008], and PWD wellness [Χ^2^ (2, *N =* 202) = 6.749, p=.034]. There were no main effects of adulthood phase on desired support for caregiving skills, caregiver wellness, or medical advice related to the PWD. Post-hoc test revealed that relative to both middle and late adulthood caregivers, early adulthood caregivers more frequently endorsed caregiver mental health as a desired support area (see Table 4 for post-hoc comparisons). Middle adulthood caregivers, relative to the two other groups, more frequently endorsed wanting support with advanced care planning. Relative to late adulthood caregivers only, early adulthood caregivers more frequently endorsed wanting support in the form of supplemental PWD care and PWD safety support. There were no post-hoc differences in desired supports between adulthood phases for PWD wellness.

#### Continuous Age Analyses

When age was examined as a continuous variable in this context, caregivers who desired caregiver mental health support, supplemental PWD care, PWD safety, and PWD wellness were significantly younger than those who did not endorse these needs (see Table 4). There were no other significant mean age differences of those who did versus did not endorse areas of support.

## Preferences for Intervention

### Overall Sample

In the overall sample, approximately half of the caregivers reported a likelihood to use support programs in the form of individual counseling (n=106, 52.5%) and a guided support group (n=111, 55%). Approximately one-third (n=68, 33.7%) said they would use a digital support program that did not involve human-to-human interaction, such as listening to podcasts, watching online videos, or using a self-guided mobile app. In terms of modality, the most commonly endorsed preferred setting for support program delivery was a combination of in-person and online (n=114, 59.7%), with fewer caregivers endorsing a desire for in-person only (n=19, 10%) and online only (n=58, 30.4%) support.

### Caregiver Adulthood Phase Analyses

#### Categorical Adulthood Phase Analyses

There were significant differences in adulthood phase and self-reported likelihood to engage in individual counseling [Χ ^2^ (2, *N* = 202) = 8.362, p = .015], guided support group counseling [Χ ^2^ (2, *N* = 202) = 7.437, p = .024], and digital support programs with no human-to-human interaction [Χ ^2^ (2, *N* = 202) = 7.654, p = .022]. Post-hoc pairwise comparisons revealed that early adulthood caregivers more frequently self-reported that they would be likely to use individual counseling and digital support programs relative to late adulthood caregivers (see Table 5). Middle adulthood caregivers were more likely than both early and late adulthood caregivers to say they would use a guided support group. There were no significant differences in preferences for support group settings (in-person, online, or hybrid) across adulthood phase groups.

**Table 5.**
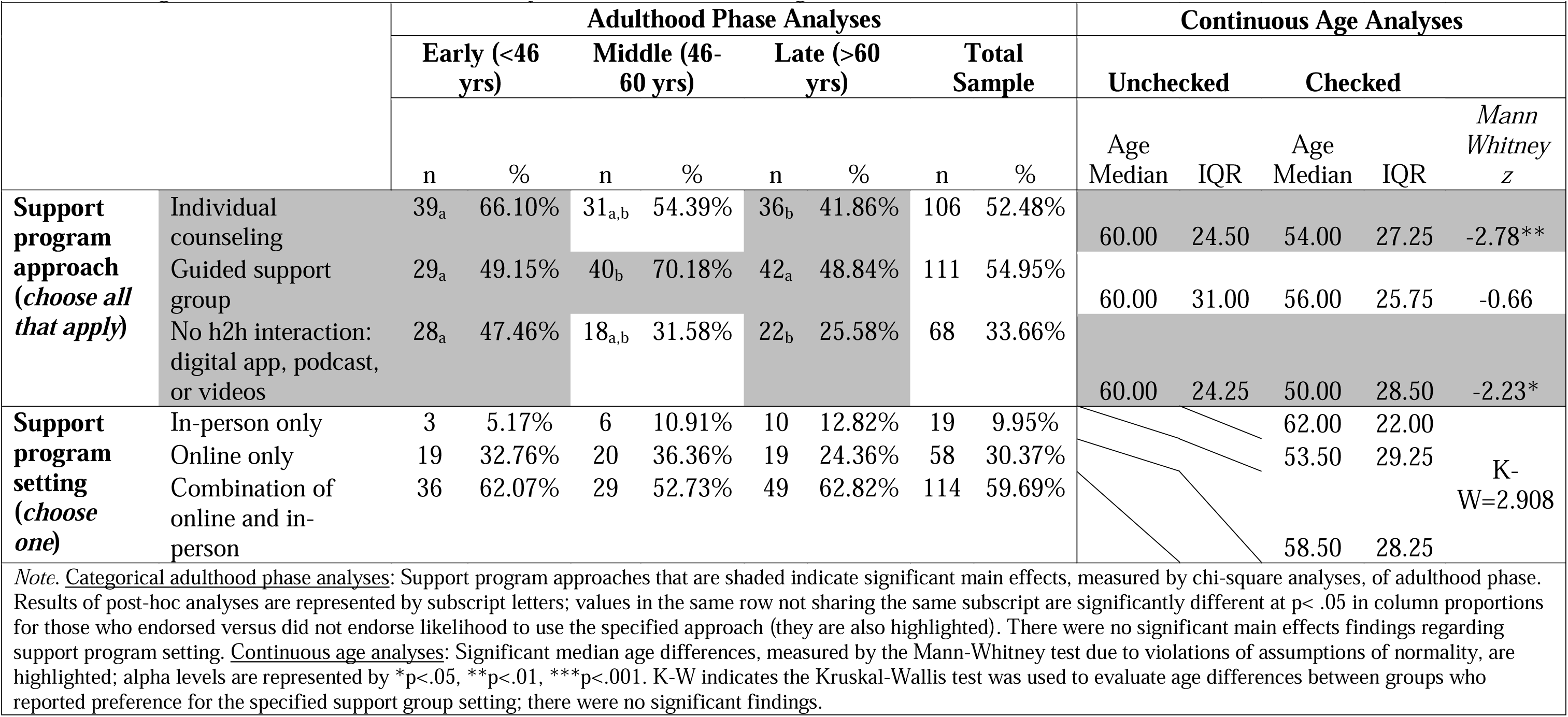
Caregiver Preferences for Intervention by Adulthood Phase and Age.

#### Continuous Age Analyses

When age was examined as a continuous variable, caregivers who were willing to engage in individual counseling and digital support programs were significantly younger than those who did not endorse these platform options. There were no significant differences in age between those who did or did not endorse likely engagement in a guided support group, nor between support program setting preference groups.

## Discussion

This cross-sectional study identified differences in mental health, perceived time pressure, support needs, and intervention preferences across adulthood phases among dementia caregivers. Caregivers in early adulthood (aged <46 years) reported higher anxiety and greater perceived time pressure than caregivers in late adulthood (aged >60), and higher time pressure than those in middle adulthood (aged 46-60). Early adulthood caregivers were also more likely than select older groups to desire additional support for their own mental health, supplemental PWD care, and PWD safety resources. In contrast, middle adulthood caregivers more frequently endorsed desired support for advanced care planning. Early adulthood caregivers further expressed greater interest in individual counseling and digitally-delivered interventions without human-to-human interaction compared to late adulthood caregivers. Collectively, these findings support hypotheses that there are distinct psychological demands, desired support areas, and service preferences across caregivers from differing adulthood phases.

Early adulthood caregivers’ heightened anxiety, time pressure, and desired support for their own mental health and caregiving relief options fit within the life course theoretical framework. This framework emphasizes the importance of the timing of role transitions, role strain that arises from competing obligations, and the accumulation of stress when significant obligations occur earlier than expected (Elder and George 2016). Early adulthood is typically marked by career building, partner establishment, and parenting, with little free time compared to later adulthood (Hutteman *et al*. 2014; Lachman *et al*. 2015). Therefore, assuming a dementia family caregiving role in this period may intensify stress and exacerbate perceptions of time scarcity. This is consistent with prior work among caregivers of adults with all chronic conditions (not just dementia) demonstrating elevated psychological distress among younger and working family caregivers (Gallagher *et al*. 2025; Kayaalp *et al*. 2021). The greater desire for mental health support, supplemental care, and safety-related resources among younger caregivers in this study aligns with evidence that adults in early and middle-adulthood prioritize interventions that reduce immediate burden and preserve role functioning across work and family domains, rather than interventions oriented toward role acceptance or meaning-making that are more common in later adulthood (Lachman *et al*. 2015).

Intervention delivery preferences further reflect cohort- and life-stage–specific constraints. Early adulthood caregivers’ greater interest in individual counseling and asynchronous digital interventions suggests a preference for individualized, flexible, private, and time-efficient formats that can be integrated into complex schedules. These preferences are consistent with broader patterns of technology adoption and expectations for on-demand services among younger cohorts of caregivers (Project Catalyst and HITLAB 2016). Together, these findings underscore that caregiving in early adulthood constitutes a developmentally distinct experience, not merely an earlier version of later-life caregiving, and support the application of life-course–informed intervention design.

Methodologically, this study highlights the utility and limitations of treating age as a continuous variable versus categorizing caregivers by adulthood phase (age groups). Some outcomes, such as time pressure, demonstrated consistent linear associations with age; however, several findings were evident only when adulthood phase was examined categorically. For example, middle adulthood caregivers uniquely endorsed greater interest in guided support groups and advanced care planning, patterns that were not captured in continuous age analyses. These results suggest that reliance on continuous age alone may obscure non-linear or phase-specific differences that are relevant for intervention tailoring. Explicit consideration of adulthood phase may therefore improve the precision of caregiver intervention design and targeting.

An additional finding of note is that caregivers across all adulthood phases expressed a clear preference for hybrid intervention delivery models that combine in-person and online delivery. This adds to a growing body of literature highlighting family caregivers’ preference for hybrid delivery options that blend in-person and online elements, as well as blending automated and human-delivered components (Blackberry *et al*. 2023; Cui *et al*. 2025; Levinson *et al*. 2020; Ramirez *et al*. 2021). Notably, preference and uptake are not synonymous and future research is warranted regarding delivery platforms that are usable and effective. Nonetheless, hybrid models may offer optimal balance between accessibility and relationship support. However, implementing hybrid services and interventions presents several challenges. A national shortage of mental health providers, particularly with caregiving burden or stress expertise, to provide individual or group services limits the scalability of support services with human-delivered elements (Applebaum and Sannes 2025). Insurance reimbursement barriers for services related to caregiving burden poses an additional constraint to models with human-delivered elements (Applebaum and Sannes 2025). Despite these challenges, our findings suggest that hybrid models warrant continued investment and evaluation.

Overall, interventions to address dementia caregiver stress, burden, and burnout may provide maximum benefit if they are optimized to meet the needs and preferences of the caregivers’ adulthood phase. For example, the results of this study would support an early adulthood caregiver intervention with components that reduce anxiety, stress, and perceived time pressure; address caregiving relief options and care-recipient safety; and use both digitally automated content and live human-to-human support as delivery methods. By contrast, a middle adulthood caregiver intervention might warrant greater emphasis on advanced care planning for the care-recipient and be delivered in group format.

There are several important limitations to this study. The cross-sectional nature of this study hampers the ability to identify causal relationships between adulthood phase and outcomes of interest. Additionally, the majority of study participants are from Virginia, which limits the generalizability of findings to national caregiving cohorts. However, Virginia is an economically and geographically diverse state, and there is wide representation of the rural/urban and socioeconomic spectrum in this study. Further, stated intervention preferences are not necessarily indicative of engagement, adherence, or uptake in real-world settings. This highlights the need for future research that rectifies these gaps.

With these limitations in mind, the present findings underscore the importance of attending to adulthood phase when assessing dementia caregivers’ mental health needs, support priorities, and intervention preferences. As the number of dementia caregivers from earlier phases of adulthood increases, interventions that explicitly account for life-stage–specific role demands, time pressure, and delivery preferences may improve relevance, engagement, and effectiveness. Future longitudinal and intervention studies are needed to test life course–informed, hybrid intervention models and to evaluate whether tailoring intervention content and delivery by adulthood phase leads to improved caregiver outcomes and more efficient use of limited support resources.

## Data Availability

All data produced in the present study are available upon reasonable request to the authors.

## Supplementary Material

### Fraud screening process

First, screening form responses were reviewed for suspicious activity such as multiple screening forms completed successively in a short amount of time with slightly adjusted names or email addresses (e.g., changing the last name by one letter). Suspicious activity was noted in the study research log. Second, potential participants were contacted for an information verification call, in which individuals were asked to verify their basic contact information and answers to several screening questions. If the individual provided discrepant information or the provided phone number had been disconnected, the suspicious activity was noted in the research log. All suspicious activity was discussed by the study team until a consensus was reached on whether or not the individual was likely to be faking an identity or providing fabricated information.

### Missing data

All participants had complete datasets except for n=1 early adulthood caregiver missing responses, and therefore total scores, for three (WHO-5, PACS, and PCS) questionnaires, n=1 middle adulthood caregiver missing four (WHO-5, Time Pressure, PACS, and PCS) questionnaires, n=7 late adulthood caregivers who were missing one (WHO-5, Time Pressure, or PACS) questionnaire, and n=2 late adulthood caregivers missing two (PACS and PCS) questionnaires. For each analysis, ≤5% of data was missing and pairwise deletion was used.

### Data cleaning

Several demographic and caregiving characteristics variables were recoded into fewer categories to facilitate interpretation of the data (for example, when subgroup sizes were n <5). The Demographics and Caregiving Characteristics form in the Supplementary Material is annotated to indicate which variables were recoded for interpretation. To reduce the size of tables and enhance readability, ‘Prefer not to answer’ responses were not included in tables. See Supplemental Material for information about missing data.

